# Long-Term Efficacy and Safety of GLP-1R Agonist and SGLT2 Inhibitor Therapy in the General Population: A 2x2 Factorial Mendelian Randomization Study

**DOI:** 10.1101/2025.07.15.25331531

**Authors:** Subbaramireddy Remala, Liming Liang, Amil M. Shah, Leo F. Buckley

## Abstract

**Introduction:** Sodium-glucose cotransporter-2 (SGLT2) inhibitors and glucagon-like peptide-1 receptor (GLP-1R) agonists reduce the risk of major adverse cardiovascular and kidney events in individuals with various cardiometabolic conditions. The long-term efficacy and safety of these therapies, especially in low- and moderate-risk populations, remain uncertain.

**Methods:** We conducted a biobank-scale analysis using genetic instruments derived from naturally occurring genetic variations in the genes encoding the targets of SGLT2 inhibitors (SLC5A2) and GLP-1R agonists (GLP1R) that associated with HbA1c levels. This Mendelian randomization study utilized data from the All of Us Research Program, which includes whole genome sequencing and electronic health records of 633,547 participants.

**Results:** Higher SGLT2 inhibitor genetic instrument scores were associated with a lower risk of heart failure (OR: 0.97, 95% CI: 0.96 to 0.99) and chronic kidney disease (OR: 0.98, 95% CI: 0.96 to 0.99). Higher GLP-1R agonist genetic instrument scores were linked to reduced risks of heart failure (OR: 0.97, 95% CI: 0.95 to 0.99), chronic kidney disease (OR: 0.96, 95% CI: 0.94 to 0.98), and coronary artery disease (OR: 0.98, 95% CI: 0.96 to 0.99). We did not detect associations between the GLP1-R agonist instrument and multiple endocrine neoplasia or medullary thyroid carcinoma. PheWAS identified associations between the SGLT2 inhibitor and GLP-1R agonist genetic instruments and a lower risk of diabetes, but no other phenotypes.

**Conclusions:** This study demonstrates the utility of biobank-scale health data for pharmacology research and suggests that, if feasible to implement in routine practice, long-term, combined primary prevention with an SGLT2 inhibitor or GLP-1R agonist would safely lower the risk of major adverse cardiovascular and kidney events in low- to moderate-risk adults.

## Introduction

Sodium-glucose cotransporter-2 (SGLT2) inhibitors and glucagon-like peptide-1 receptor (GLP-1R) agonists lower the risk of major adverse cardiovascular and kidney events across a wide range of individuals with cardiovascular, kidney and metabolic conditions.^1^ Societal guidelines recommend the routine use of SGLT2 inhibitors and GLP-1R agonists in patients with type 2 diabetes^2^, heart failure^3^ and chronic kidney disease^4^.

Several questions about SGLT2 inhibitors and GLP-1R agonists remain unanswered. Follow-up in the landmark SGLT2 inhibitor and GLP-1R agonist clinical trials ended after 2-3 years, creating uncertainty about the efficacy and safety of long-term exposure to these novel mechanisms of action. Additionally, landmark clinical trials focused enrollment on high-risk individuals likely to experience an event within a few years of enrollment and the efficacy of SGLT2 inhibitors and GLP-1R agonists for the primary prevention of cardiovascular and kidney events in low- and moderate-risk individuals, who account for a larger number of clinical events^5^, is unclear. Based on post-hoc subgroup analyses from GLP-1R agonist and SGLT2 inhibitor clinical trials in type 2 diabetes, combination therapy appears safe and efficacious.^6,7^ Data on combination GLP-1R agonist and SGLT2 inhibitor therapy outside of type 2 diabetes is lacking. Last, the effects of GLP-1R agonists on medullary thyroid cancer in mice has led to a Black Box Warning on the use of these agents in patients with a personal or family history of medullary thyroid cancer or multiple endocrine neoplasia type 2, but data from humans are limited to small samples sizes due to the rarity of these conditions.

Addressing these questions using randomized controlled trials would require considerable resources to enroll large sample sizes and enable long-term follow-up. Therefore, we conducted a biobank-scale analysis using naturally occurring genetic variation in the genes encoding the targets of SGLT2 inhibitors (SLC5A2) and GLP-1R agonists (GLP1R) to create genetic instruments that mimic the pharmacologic effects of lifelong exposure to these therapies. Under the assumptions of a Mendelian randomization analysis framework, these genetic instruments are free from reverse confounding and other potential sources of bias found in observational studies. Our aim was to estimate the long-term efficacy and safety of SGLT2 inhibitor and GLP-1R agonist therapy in a general population.

## Methods

### All of Us Research Program Design

The All of Us Research Program (All of Us) aims to establish a biobank of one million participants across the United States.^8^ All of Us builds upon and complements existing biobanks through its size and enrollment of an expansive population representative of all individuals in the United States. All of Us participants provide data from an in-person study visit and can participate in optional ancillary studies that collect data from wearables, surveys, and other modalities. In addition, participants from select enrollment centers can opt to provide access and linkage to their electronic health records. We used the All of Us Curated Data Repository v8 (C2024Q3R5, release date February 2025) for this analysis, which included 633,547 participants who enrolled and provided written informed consent before October 1, 2023.

### Study Population

The study population included All of Us participants with available electronic health record data and short-read whole genome sequencing data.^9^ We created a training cohort to develop the SGLT2 inhibitor and GLP-1R agonist genetic instruments and different outcomes cohorts to test their associations with cardiometabolic traits and incident clinical outcomes (**Supplemental Figure 1**).

The training cohort included the subset of the overall All of Us population with electronic health record and whole genome sequencing data who had at least one hemoglobin A1c (HbA1c) value (winsorized to values less than 20%) in the linked electronic health record data that was measured prior to the first occurrence of a diabetes diagnosis or prescription of glucose-lowering therapy (**Supplemental Table 1**). Participants who were less than 18 years old at the time of HbA1c measurement were excluded.

We tested the associations between the genetic instruments and cardiometabolic traits and clinical outcomes in dedicated outcomes cohorts. The cardiometabolic trait cohorts included All of Us participants with whole genome sequencing data and physical measurement data (for blood pressure, body mass index, and waist-to-hip ratio) or electronic health record-derived hematocrit and hemoglobin levels. The clinical outcomes cohort included All of Us participants with whole genome sequencing data and linked electronic health record data.

The phenome-wide association study (PheWAS) cohort included All of Us participants with short-read whole genome sequencing data and linked electronic health record data.

### GLP-1R Agonist and SGLT2 Inhibitor Genetic Instruments Construction

We built the GLP-1R agonist and SGLT2 inhibitor genetic instruments from single nucleotide polymorphisms and insertion-deletion variants that reside within exonic regions 200 kilobases (kb) upstream and downstream of the GLP1R and SLC5A2 genes. A total of 10,740 GLP1R variants and 17,847 SLC5A2 variants were eligible for inclusion in the genetic instruments.

We used elastic net regression models with forced adjustment for age, sex, and 16 principal components of genetic ancestry to identify variants that associated with HbA1c levels. We defined the effect allele as the variant associated with lower HbA1c levels. We used 10-fold cross-validation to select the GLP-1R agonist and SGLT2 inhibitor models that minimized the mean-squared error.

Next, we pruned the selected HbA1c-associated variants in GLP-1R agonist and SGLT2 inhibitor models using a linkage disequilibrium threshold of r^2^ < 0.3, 200 kb window size and 50 kb step size. The GLP-1R agonist and SGLT2 inhibitor genetic instruments were created as linear combinations of the number of HbA1c-associated alleles, weighted by the beta-coefficients for the effect allele association with HbA1c.

### Cardiometabolic Traits

We selected the following traits known to be modified by GLP-1R agonist or SGLT2i to confirm the validity of the instruments and assess their strength: body mass index, systolic blood pressure, diastolic blood pressure, hemoglobin, hematocrit and waist-to-hip ratio. Body weight, systolic blood pressure, diastolic blood pressure and waist-to-hip ratio were obtained at the in-person study visit according to standardized procedures.^8^ Hemoglobin and hematocrit values were extracted from linked electronic health record data.

### Clinical Outcomes

The primary clinical outcomes of interest were heart failure (HF), chronic kidney disease (CKD) and coronary artery disease (CAD). Electronic health record International Classification of Diseases (ICD) codes were mapped onto Systematized Nomenclature of Medicine - Clinical Terms (SNOMED) codes in the All of Us data. Participants were considered to have experienced a clinical outcome of interest if they had at least two distinct SNOMED recordings in their electronic health record data (**Supplemental Table 2)**. We included as safety outcomes the composite of multiple endocrine neoplasia or medullary thyroid carcinoma and any thyroid cancer defined using SNOMED codes (**Supplemental Table 3**).

We conducted a PheWAS to test the associations between the GLP-1R agonist and SGLT2 inhibitor genetic instruments and a wide range of phenotypes. The PheWAS allowed us to test the associations between the GLP-1R agonist and SGLT2 inhibitor instruments and known outcomes as a positive control, assess the associations between these genetic instruments and potential adverse effects and identify novel hypothesis-generating associations for potential drug repurposing. ICD codes in the linked All of Us electronic health record data were aggregated into 1,843 phenotypes as described previously.^10^ Two ICD codes were required for a positive phenotype. Sex-specific phenotypes were analyzed only in All of Us participants with the appropriate self-reported sex at birth.

### Statistical Analysis

Characteristics of the training and testing cohorts were summarized using means (standard deviation) and numbers (%). To confirm that the GLP-1R agonist and SGLT2 inhibitor genetic instruments mimicked the physiologic effects of the drugs, we used linear regression models to test the associations between the GLP-1R agonist and SGLT2i genetic instruments and body mass index, systolic blood pressure, diastolic blood pressure, hemoglobin, hematocrit and waist-to-hip ratio among participants in the cardiometabolic traits testing cohorts. We calculated the F-statistic for models of each cardiometabolic trait with only the genetic instrument score to quantify the strength of the instrument.^11^

We used logistic regression models to test the associations between the GLP-1R agonist and SGLT2 inhibitor genetic instruments and the odds of HF, CKD and CAD. All models were adjusted for age, sex, the first 16 genetic ancestry principal components and an indicator variable for the availability of an HbA1c value. We tested whether the association between the GLP-1R agonist instrument and the odds of HF, CKD and CAD differed according to the SGLT2 inhibitor instrument (and the converse) by adding a multiplicative interaction term in the logistic regression models. The PheWAS was conducted using logistic regression models adjusted for age, sex, the first 16 principal components analysis and an A1c indicator variable. For the genetic instrument associations with cardiometabolic traits and clinical outcomes, a P-value or P-interaction < 0.05 was considered statistically significant since these associations were prespecified based on prior knowledge. A Bonferroni-corrected P-value < 0.05 was considered statistically significant in the PheWAS analysis.

## Results

### Participant Characteristics

The genetic instrument training cohort included 124,137 participants with a mean (SD) age of 52 (15) years (48,612 [39%] men, 73,456 [59%] women, and 2,069 [2%] others) (**Supplemental Table 4**). The outcomes cohorts included 317,964 individuals with a mean (SD) age of 56 (17) years (122,069 [38%] men, 192,551 [61%] women, and 3,344 [1%] others) (**Supplemental Table 4**). The cardiometabolic trait cohorts included between 209,330 and 296,769 individuals (**Supplemental Table 5**).

### Cardiometabolic Traits

The SGLT2 inhibitor genetic instrument included 31 variants with a median beta coefficient of −0.0035% (25^th^, 75^th^ percentiles: −0.0009% to −0.0025%) HbA1c per 1-additional effect allele and a median minor allele frequency of 0.062 (25^th^, 75^th^ percentiles: 0.025 to 0.203) (**Supplemental Table 6**). The GLP-1R agonist genetic instrument included 23 variants with a median (25^th^, 75^th^ percentile) beta coefficient of −0.0069% (−0.014% to −0.003%) HbA1c per 1-additional effect allele and a median (25^th^, 75^th^ percentile) minor allele frequency of 0.130 (0.047 to 0.397) (**Supplemental Table 7**). Individuals with an SGLT2 inhibitor or GLP-1R agonist genetic instrument score above the median (corresponding to lower HbA1c) had a similar mean age and a similar proportion of women and men as those with scores below the median (**Table 2**).

**Table 1.** Participants characteristics according to SGLT2 inhibitor and GLP-1R agonist genetic instrument score.

|  | SGLT2 Inhibitor Instrument |  | GLP-1R Agonist Instrument |  |
| --- | --- | --- | --- | --- |
|  | Below Median | Above Median | Below Median | Above Median |
| Age, years | 56 (17) | 57 (17) | 56 (17) | 57 (17) |
| Men | 60,754<br>(38%) | 61,315<br>(39%) | 61,007<br>(38%) | 61,062<br>(38%) |
| Women | 96,764<br>(61%) | 95,787<br>(60%) | 96,044<br>(60%) | 96,507<br>(61%) |
| Other | 1,627<br>(1%) | 1,717<br>(1%) | 1932<br>(2%) | 1414<br>(1%) |
| Hypertension | 80,241<br>(50%) | 83,151<br>(52%) | 83,935<br>(53%) | 79,457<br>(50%) |
| Diabetes | 50,722<br>(32%) | 51,402<br>(32%) | 52,366<br>(33%) | 49,758<br>(31%) |
| Heart Failure | 9,713<br>(6%) | 10,154<br>(6%) | 10,508<br>(7%) | 9,359<br>(6%) |
| Coronary Artery Disease | 13,794<br>(9%) | 14,138<br>(9%) | 13,483<br>(8%) | 14,449<br>(9%) |
| Chronic Kidney Disease | 11,257<br>(7%) | 11,665<br>(7%) | 12,020<br>(8%) | 10,902<br>(7%) |

**Table 2.** GLP-1Ra and SGLT2i genetic instrument associations with multiple endocrine neoplasia and medullary thyroid carcinoma.

| Outcome | Number of Events (%) | Instrument | OR (95% CI) | P-Value |
| --- | --- | --- | --- | --- |
| Multiple Endocrine Neoplasia or Medullary Thyroid Carcinoma | 75 (0.02%) | SGLT2i | 0.93 (0.82-1.19) | 0.46 |
|  |  | GLP1Ra | 0.93 (0.68-1.21) | 0.62 |
| Thyroid Cancer of Any Type | 2,647 (0.83%) | SGLT2i | 1.00 (0.96-1.05) | 0.76 |
|  |  | GLP1Ra | 1.01 (0.96-1.05) | 0.61 |

A 1-unit higher SGLT2 inhibitor genetic instrument was associated with a significantly lower HbA1c level (beta coefficient [95% CI]: −0.052 [−0.059 to −0.045]; P<0.001, F-stat = 219.3). A 1-unit higher GLP-1R agonist genetic instrument was associated with a significantly lower HbA1c level (beta coefficient [95% CI]: −0.059 [−0.066 to −0.052]; P<0.001, F-stat = 288.7). A higher SGLT2 inhibitor genetic instrument score was associated with significantly lower body mass index, lower waist-to-hip ratio, lower diastolic blood pressure, higher hematocrit and higher hemoglobin (**Figure 1A**). A higher GLP-1R agonist score associated with lower body mass index and lower systolic and diastolic blood pressure, but not with hemoglobin or hematocrit (**Figure 1B**).

**Figure 1.**
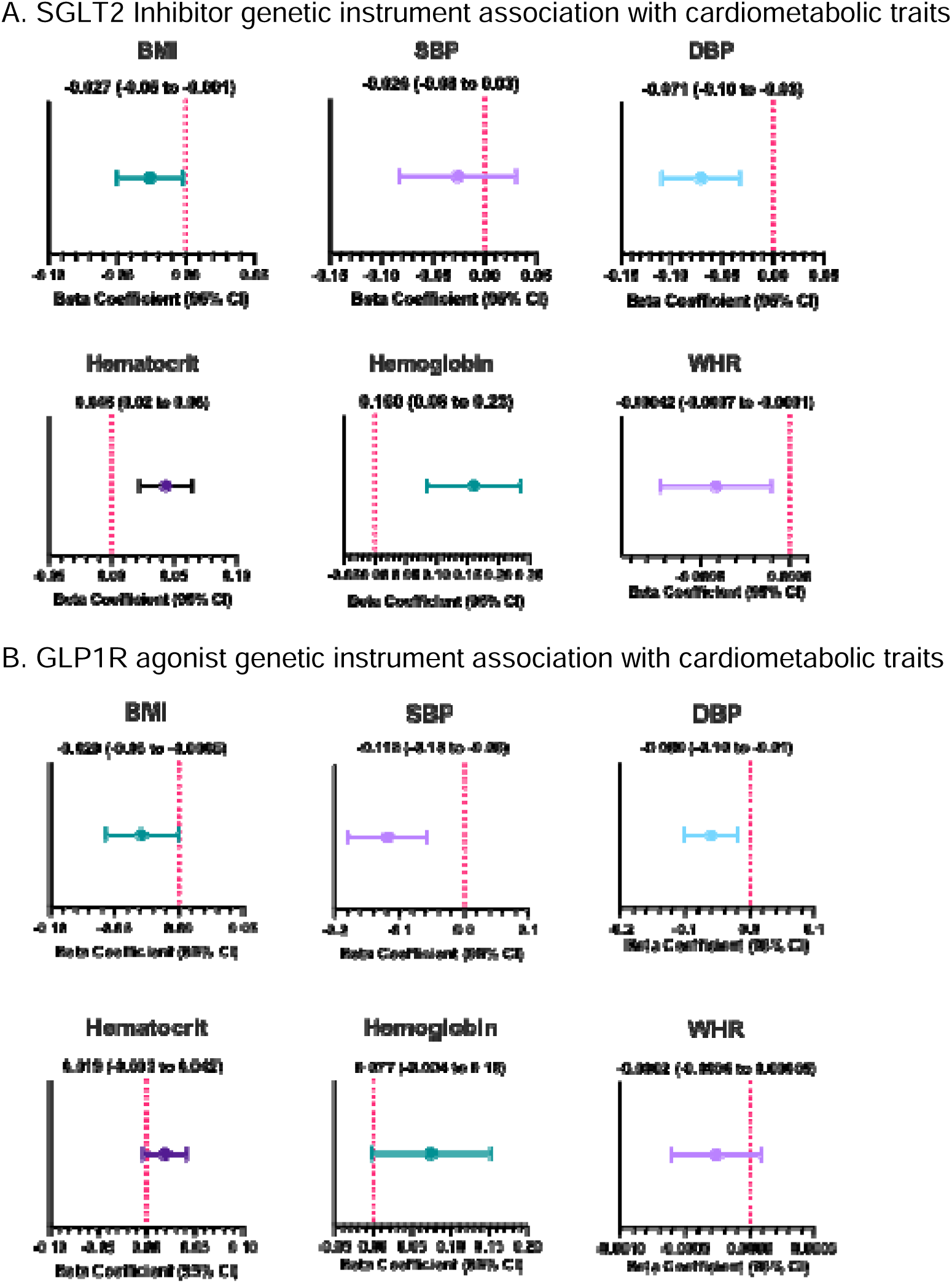
SGLT2 inhibitor and GLP-1R agonist genetic instrument associations with cardiometabolic traits BMI = body mass index; DBP = diastolic blood pressure; SBP = systolic blood pressure; WHR = waist-to-hip ratio. <u>Caption</u>: Each panel forest plot displays an effect size and 95% confidence intervals from linear regression models evaluating the relationship between the SGLT2 inhibitor genetic instrument (panel A) or the GLP-1R agonist genetic instrument (panel B) and various cardiometabolic traits.

### Efficacy Outcomes

In the clinical outcomes cohort, there were 19,867 (6%) incident HF events, 22,922 (7%) incident CKD events, and 27,932 (9%) incident CAD events. A higher SGLT2 inhibitor genetic instrument score (mimicking greater SGLT2 inhibition) was associated with a significantly lower risk of incident HF (OR: 0.97, 95% CI: 0.96 to 0.99, P-value = 0.004) and incident CKD (OR: 0.98, 95% CI: 0.96 to 0.99, P-value = 0.01) (**Figure 2A**). A higher GLP-1R agonist instrument (indicating greater GLP-1R agonism) was associated with a lower risk of incident HF (OR: 0.97, 95% CI: 0.95 to 0.99, P-value = 0.01), incident CKD (OR: 0.96, 95% CI: 0.94 to 0.98, P-value <0.001) and incident CAD (OR: 0.98, 95% CI: 0.96 to 0.99, P-value = 0.01), (**Figure 2B**).

**Figure 2.**
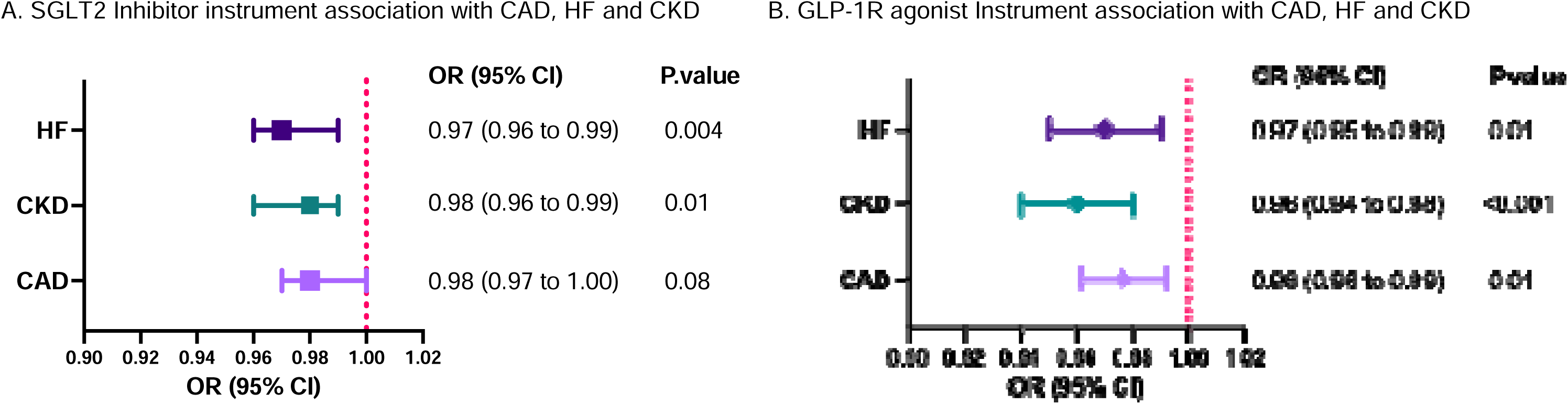
SGLT2 inhibitor and GLP-1R agonist genetic instrument associations with clinical outcomes CAD = coronary artery disease; CI = confidence interval; CKD = chronic kidney disease; HF = heart failure; OR = odds ratio <u>Caption</u>: Forest plot displays an odd ratio and 95% confidence intervals from logistic regression models evaluating the relationship between the SGLT2 inhibitor (panel A) or GLP-1R agonist (panel B) genetic instruments and incident cardiometabolic outcomes.

The associations between the GLP-1R agonist genetic instrument and incident CAD (P-interaction = 0.92) and incident HF (P-interaction = 0.61) were not significantly modified by the SGLT2 inhibitor genetic instrument (**Supplemental Table 8**). Similarly, the associations between the SGLT2 inhibitor genetic instrument and incident CAD (P-interaction = 0.18) and incident HF (P-interaction = 0.79) were not modified by the GLP-1R agonist instrument (**Supplemental Table 8**). We detected a significant interaction between the GLP-1R agonist instrument and the SGLT2 inhibitor instrument for incident CKD. The associations between the GLP-1R agonist instrument was stronger in the subgroup with an SGLT2 inhibitor genetic instrument score below the median (OR [95% CI]: 0.95 [0.93-0.97]) compared to above the median (OR [95% CI]: 0.98 [0.96-1.01]) (P-interaction = 0.032). A similar trend was observed for the association between the SGLT2 inhibitor instrument across levels of the GLP-1R agonist instrument (P-interaction = 0.079) (**Supplemental Table 8**).

### Safety Outcomes

There were 75 cases of multiple endocrine neoplasia (n=70) or medullary thyroid carcinoma (n=5) and 2,647 cases of any thyroid cancer. The GLP-1R agonist genetic instrument had no significant associations with the composite of multiple endocrine neoplasia or medullary thyroid carcinoma or any thyroid cancer **(Table 2).**

PheWAS identified associations between the SGLT2 inhibitor and GLP-1R agonist genetic instrument and a lower risk of diabetes-related Phecodes (**Supplemental Table 9**). There were no associations between the SGLT2 inhibitor or GLP-1R agonist genetic instruments and other phenotypes in the PheWAS (**Figure 3**).

**Figure 3.**
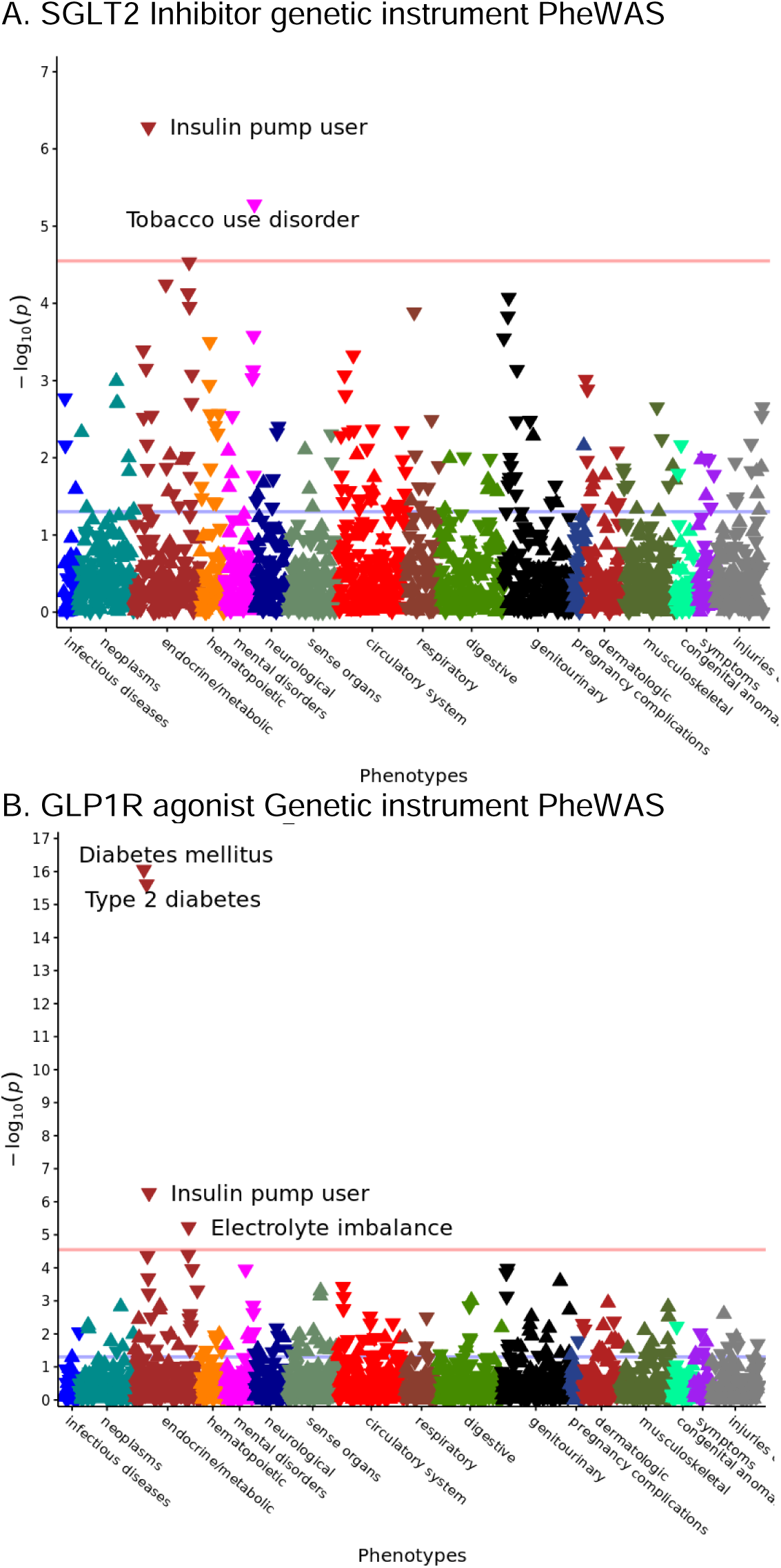
Phenome-wide association studies for the SGLT2 inhibitor and GLP-1R agonist genetic instruments <u>Caption</u>: The vertical axis shows the -log_10_ Bonferroni-adjusted P-value for the specified phenotype on the horizontal axis. Each point represents an association result. Points above the horizontal red line are statistically significant.

## Discussion

Using whole genome sequencing data from 124,137 participants in the All of Us Research Program, we built genetic instruments to mimic the pharmacologic effects of SGLT2 inhibitors and GLP-1R agonists, two cardiometabolic therapies with established efficacy and safety in individuals at high risk for major adverse cardiovascular and kidney events. We used these two genetic instruments to address outstanding clinical questions about SGLT2 inhibitors and GLP-1R agonists that cannot be investigated using other approaches. We found that higher SGLT2i genetic instrument scores were associated with a lower lifetime risk of heart failure and chronic kidney disease, but not coronary artery disease, while higher GLP-1R agonist scores were associated with lower lifetime risks of coronary artery disease, heart failure, and chronic kidney disease. Moreover, the associations between the SGLT2 inhibitor and GLP-1R agonist instruments and cardio-kidney outcomes were independent of each other. Last, our phenome-wide association study did not identify any new safety signals for either SGLT2 inhibitors or GLP-1R agonists and or any evidence to support an association between GLP-1R agonists and medullary thyroid carcinoma or multiple endocrine neoplasia. Altogether, these findings demonstrate the utility of biobank-scale health data for pharmacology research and suggest that, if it were feasible to implement in routine practice, long-term, combined primary prevention with an SGLT2 inhibitor or GLP-1R agonist may safely lower the risk of major adverse cardiovascular and kidney events in low- to moderate-risk adults.

The use of observational study designs to infer causal relationships related to pharmacology has attracted considerable attention. Comparative effectiveness and target trial emulations use increasingly large cohorts of routinely collected health data to compare the effects of two approved drug therapies on clinical outcomes of interest.^12^ These approaches can lead to valid findings depending upon the availability of the data needed to faithfully replicate the target trial.^13^ Alternatively, pioneering studies on drug target genetic instruments used summary statistics or individual-level meta-analysis of well-phenotyped, small- and moderate-sized cohorts.^14,15^ An advantage of using genetic instruments to mimic drug target effects is the potential to create instruments for targets without approved therapies being used in clinical practice. The growing number and size of health system-specific and publicly available biobanks with genomic data has enabled improved genomic-based observational drug target studies.^16–19^ We demonstrate the feasibility of creating strong, valid drug target genetic instruments from biobank-scale whole genome sequencing data linked to electronic health records. Our large sample size and use of whole genome sequencing allowed us to aggregate the effects of multiple, common variants with small effect sizes. This particular aspect was integral to the creation of the SGLT2 inhibitor instrument as genetic variants in SLC5A2 impair SGLT2 function and leads to glucosuria, but no other observable phenotype.^20^ A cohort enriched with rare variants and larger effect sizes would likely have the long-term follow-up needed to accrue sufficient clinical events for a well-powered analysis. The volume of linked electronic health record data also facilitated phenome-wide association studies to detect safety signals and even the investigation of rare conditions such as medullary thyroid carcinoma. These genetic instruments can be used to investigate the potential effects of established drug therapies on novel outcomes, rule out large safety signals, and stratify individuals according to predicted drug response. The long-term safety data have implications for the design of novel therapies with long-term effects, such as RNA interference. The approach described in this study may be adapted to inform the long-term safety of other therapeutic targets.

Several aspects of our study design support the validity of using genetic instruments to mimic the pharmacologic effects of SGLT2 inhibitors and GLP-1R agonists. We included only exonic genetic variants within the genomic region encoding the protein drug target to minimize the potential for horizontal pleiotropy and used regularized regression and cross-validation to create parsimonious models.^21^ In addition, we recreated associations between the genetic instruments and cardiometabolic traits known to be modified by the corresponding drugs, such as the effects of SGLT2 inhibitors on hemoglobin levels and GLP-1R agonists on body weight, as positive controls. Last, the pattern of associations between the genetic instruments and clinical outcomes agrees with the known effects of SGLT2 inhibitors on HF and CKD, but less so on CAD, and GLP-1R agonists on HF, CKD and CAD.

Landmark clinical trials have established the efficacy and safety of SGLT2 inhibitors and GLP-1R agonists in patients with type 2 diabetes, as well as in patients with heart failure with reduced or preserved ejection fraction or chronic kidney disease irrespective of diabetes status.^22^ For numerous reasons, these trials were enriched with individuals at high short-term risk. A large number of individuals in the general population, however, have risk factors for cardiovascular and kidney disease but do not meet trial eligibility criteria for an SGLT2 inhibitor or GLP-1R agonist^5^. Primary prevention clinical trials face numerous challenges, such as low event rates, long-term adherence and follow-up, and cost. Thus, strong evidence is needed to justify an investment in a primary prevention clinical trial. Our findings, derived from an analysis of 317,964 individuals in the general population, suggest that the established efficacy and safety of SGLT2i and GLP-1R agonists would extend to a broader population at moderate or even low short-term risk for major adverse cardiovascular and kidney events. Additional evidence, such as studies in other populations or studies that design the genetic instruments with a different approach, would further strengthen the argument for primary prevention clinical trials with SGLT2 inhibitors and GLP-1R agonists. The feasibility of such trials and the potential implementation of a new primary prevention therapy requires careful thought and planning.

Our results offer additional insights into the clinical effects of combination therapy with both an SGLT2 inhibitor and a GLP-1R agonist. Two meta-analyses that combined post-hoc subgroup results from type 2 diabetes clinical trials found no evidence of an interaction between SGLT2 inhibitors and GLP-1R agonists and their effects on cardio-kidney outcomes.^6,7^ Due to the limited use of combined therapy during the enrollment periods of those trials, however, the number of combined SGLT2 inhibitor and GLP-1R agonist users (999 and 1,660 in the SGLT2 inhibitor and GLP-1R agonist meta-analyses, respectively) and the number of events (a total of 400 major adverse cardiovascular events, 214 heart failure hospitalization events and 193 CKD progression events across both meta-analyses) available for analysis were low, a limitation of subgroup analysis.^23^ Our results complement those meta-analyses by including participants without type 2 diabetes and using a large sample size with 19,867 incident HF events, 22,922 incident CKD events, and 27,932 incident CAD events. The available evidence indicates that SGLT2 inhibitors and GLP-1R agonists provide complementary benefits. Thus, clinicians should consider the use of combination therapy in individuals with dual indications. The lack of evidence of an interaction between the effects of these two drugs in our genetic analyses and the previously published post-hoc randomized clinical trial analyses argues against a synergistic effect of combination therapy.

Our phenome-wide association study found no new safety signals associated with lifelong exposure to greater GLP-1R agonism or greater SGLT2 inhibition. We also assessed the association between the GLP-1R genetic instrument and thyroid cancer risk. In mice, which express higher levels of GLP-1R on thyroid C-cells than humans, GLP-1R agonists have been associated with the development of medullary thyroid carcinoma.^24^ The United States Food and Drug Administration requires Black Box Warnings that contraindicate the use of GLP-1R agonists in patients with a personal or family history of medullary thyroid carcinoma or multiple endocrine neoplasia type 2, a precursor to medullary thyroid carcinoma. A systematic review and meta-analysis of GLP-1R agonist randomized clinical trials, which included 86 thyroid cancer events of any type (4 medullary thyroid cancer cases), median follow-up of 53 weeks, 46,228 participants receiving GLP-1R agonist treatment and 38,399 participants receiving control treatment, found a significantly higher risk with GLP-1R agonist treatment for any thyroid cancer (odds ratio [95% confidence interval]: 1.52 [1.01-2.29]; number-needed-to-harm: 1,349), but not for medullary thyroid carcinoma specifically (3 events vs. 1 event; odds ratio [95% confidence interval]: 1.44 [0.23-9.16]).^24^ A cohort study using routinely collected clinical data (mean follow-up 3.9 years) did not find an association between GLP-1R agonist use and either thyroid cancer (76 total cases) or medullary thyroid cancer (15 total cases).^25^ Our analysis, which reflects lifelong exposure to greater GLP-1R activity and included significantly more cases than prior studies (75 cases of medullary thyroid carcinoma or multiple endocrine neoplasia and 2,647 cases of thyroid cancer of any type) further strengthens the evidence base that GLP-1R agonists do not have clinically meaningful effects on thyroid cancer risk in humans.

Our study has certain limitations. We trained the genetic instrument models on HbA1c levels, which was the original primary pharmacologic effect of SGLT2 inhibitors and GLP-1R agonists but may not be the most representative trait for their overall clinical effects. Our instruments did exhibit strong associations with other known effects of SGLT2 inhibitors and GLP-1R agonists, which suggests that the instruments captured much of the important variation. We did not investigate the molecular mechanisms through which these variants (or variants in linkage disequilibrium) influence cardiometabolic traits. Targeting cis-genetic variants in exons within the GLP1R and SLC5A2 regions increased our *a priori* confidence in the genetic instruments but may miss important trans-acting variants.

## Conclusions

Drug-mimicking genetic instruments can address important clinical and translational research questions. The associations between SGLT2 inhibitor and GLP-1R agonist genetic instruments and cardio-kidney outcomes suggest the potential for these agents to be used as primary prevention in low- and moderate-risk individuals.

## Data Availability

All data produced are available online at allofus.nih.gov.

