## Supplemental Materials for "Long-Term Efficacy and Safety of GLP-1R Agonist and SGLT2 Inhibitor Therapy in the General Population: A 2x2 Factorial Mendelian Randomization Study"

**Supplemental Material**

**Supplemental** **Figure 1**. Study Workflow

EHR = electronic health record, srWGS = short-read whole genome sequence, LD = linkage disequilibrium, BMI = body mass index, SBP = systolic blood pressure, DBP = diastolic blood pressure, Whr = waist to hip circumference ratio, CAD = coronary artery disease, HF = Heart Failure, CKD = chronic kidney disease.

Caption**:** Comprehensive workflow outlining the srWGS and linked EHR integration to develop genetic instrument and association analysis.

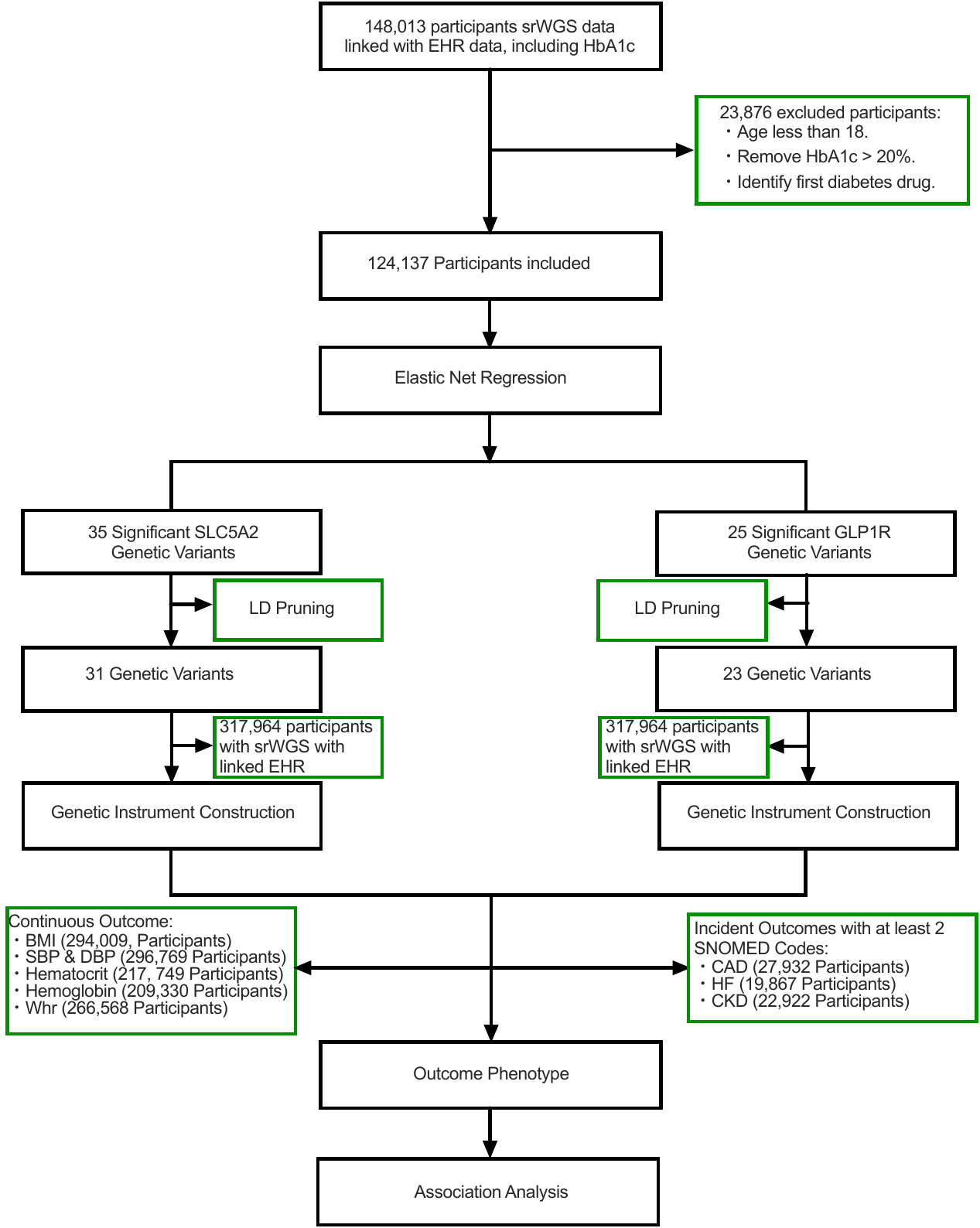

**Supplemental Table 1.** Glucose-lowering drugs excluded from the training cohort

| **Glucose-Lowering Drug Names** | **Diabetes SNOMED Codes** |
| --- | --- |
| Acarbose, Actos, Adlyxin, Albiglutide, Alogliptin, Amaryl, Avandia, Bydureon, Byetta, Dulaglutide, Exenatide, Fortamet, Glibenclamide, Glimepiride, Glipizide, Glucophage, Glucotrol, Glumetza, Glyburide, Glyset, Glynase, Insulin, Januvia, Liraglutide, Lixisenatide, Metformin, Miglitol, Nateglinide, Nesina, Onglyza, Ozempic, Pioglitazone, Precose, Pramlintide, Prandin, Repaglinide, Riomet, Rosiglitazone, Rybelsus, Saxagliptin, Semaglutide, Sitagliptin, Starlix, Symlin, Tanzeum, Trulicity, Victoza |  |

**Supplemental Table 2**. SNOMED Codes for each clinical outcome.

| **Clinical Outcome** | **SNOMED Codes** |
| --- | --- |
| Coronary Artery Disease | 401303003, 15713081000119108, 15713121000119104, 285981000119103, 703213009, 703164000, 78741000119103, 394659003, 413439005, 57054005, 17531000119105, 54329005, 70211005, 59063002, 73795002, 65547006, 76593002, 58612006, 79009004, 307140009, 401314000, 70422006, 791000119109, 15960141000119102, 59021001, 194828000, 41334000, 85284003, 89323001, 442421004, 285141000119106, 285151000119108, 442224005, 429673002, 444855007, 442240008, 724431008, 451041000124103, 371807002, 92517006, 445512009, 413838009, 117051000119103, 53741008, 139011000119104, 408546009, 233970002, 421327009, 443502000, 251024009, 697976003, 63739005, 194821006, 194843003, 300995000, 414545008, 123641001, 371804009, 451361000124102, 371803003, 22298006, 16837681000119104, 414795007, 233821000, 314207007, 719678003, 233840006, 1755008, 314116003, 233846000, 4557003, 87343002, 429245005, 315025001, 371805005, 233823002, 194842008, 233819005, 713405002, 703211006, 194857001, 194858006, 703360004, 233817007, 194802003, 429559004, 15960061000119102. |
| Heart Failure | 53931000119109, 10633002, 49584005, 15964661000119102, 443343001, 698296002, 56675007, 7421000175106, 722095005, 153951000119103, 443344007, 7401000175100, 16838951000119100, 443253003, 80479009, 359617009, 443254009, 194767001, 92506005, 71892000, 89819002, 60856006, 445236007, 153941000119100, 88805009, 79955004, 441530006, 48447003, 7411000175102, 111283005, 66989003, 10335000, 441481004, 42343007, 426263006, 426611007, 717840005,15629591000119104, 67441000119101, 15629541000119106, 23341000119109, 82523003, 83291003, 195111005, 424404003, 418304008, 16253001, 96311000119109, 84114007, 233924009, 788950000, 446221000, 703272007, 703273002, 10091002, 15781000119107, 194779001, 194781004, 5148006, 46113002, 45650007, 85232009, 83105008, 62377009, 314206003, 44313006, 367363000, 15964701000119108, 698594003, 417996009, 15629741000119102. |
| Chronic Kidney Disease | 236433006, 691421000119108, 707324008, 49708008, 709044004, 104931000119100, 96441000119101, 771000119108, 713313000, 431855005, 431856006, 129181000119109, 90731000119103, 741000119101, 433144002, 129171000119106, 90741000119107, 731000119105, 700378005, 700379002, 431857002, 129151000119102, 721000119107, 433146000, 129161000119100, 90761000119106, 714152005, 714153000, 90688005, 723190009, 111411000119103, 90791000119104, 236435004, 236436003, 46177005, 71421000119105, 96731000119100, 96721000119103, 96711000119105, 8501000119104, 285831000119108, 285881000119109, 285841000119104, 118781000119108, 10757401000119104,10757481000119108, 16726004 |

| **Clinical Outcome** | **SNOMED Codes** |
| --- | --- |
| Multiple endocrine neoplasia and medullary thyroid carcinoma | 30664006, 61808009, 61530001, 255032005 |
| Thyroid cancer of any type | 92767001, 448216007, 255028004, 423158009, 314953008, 278051002, 363478007, 255032005, 255030002, 255029007, 717734005, 94098005, 724552002 |

**Supplemental Table 3**. SNOMED Codes for Thyroid Cancers

**Supplemental Table 4.** Participant characteristics in the training and clinical outcomes cohorts.

SD = standard deviation

| **Cohort** | **N** | **Age, Mean ± SD** | **Male, n (%)** | **Female, n (%)** | **Other, n (%)** | **Diabetes, n (%)** | **Hypertension, n (%)** |
| --- | --- | --- | --- | --- | --- | --- | --- |
| Genetic Instrument Training | 124,137 | 52 ± 15 | 48,612  (39%) | 73,456  (59%) | 2,069 (2%) | 57,528 (46%) | 80,053 (64%) |
| Clinical Outcomes | 317,964 | 56 ± 17 | 122,069 (38%) | 192,551 (61%) | 3,344 (1%) | 102,124 (32%) | 163,392 (51%) |

**Supplemental Table 5**. Participant and genetic instrument characteristics in the cardiometabolic trait cohorts

NS = not significant; SD = standard deviation

| **Phenotype** | **N** | **Age, Mean ± SD** | **Male** | **Female** | **Other** | **SGLT2i Beta Coefficient (95% CI)** | **SGLT2i F-statistic** | **GLP-1Ra Beta Coefficient (95% CI)** | **GLP-1Ra F-statistic** |
| --- | --- | --- | --- | --- | --- | --- | --- | --- | --- |
| Body Mass Index | 294,009 | 57 ± 17 | 114,109 (39%) | 176,724 (60%) | 3,176 (1%) | -0.027 (-0.05 to  -0.001) | 4.5 | -0.029 (-0.05 to  -0.0006) | 1118 |
| Systolic Blood Pressure | 296,769 | 56 ± 17 | 114,334 (39%) | 179,284 (60%) | 3,151 (1%) | -0.026 (-0.08 to 0.03) | NS | -0.118 (-0.18 to  -0.05) | 835.2 |
| Diastolic Blood Pressure | 296,769 | 56 ± 17 | 114,334 (39%) | 179,284 (60%) | 3,151 (1%) | -0.071 (-0.10 to  -0.03) | 81 | -0.060 (-0.10 to  -0.01 | 967.4 |
| Hematocrit | 217,749 | 58 ± 17 | 79,955  (37%) | 135,624 (62%) | 2,170  (1%) | 0.045 (0.02 to 0.06) | 8.9 | 0.019 (-0.003 to  0.042) | NS |
| Hemoglobin | 209,330 | 58 ± 17 | 76,991 (37%) | 130,222  (62%) | 2117 (1%) | 0.160 (0.08 to 0.23) | 0.54 | 0.077 (-0.004 to  0.15) | NS |
| Waist-to-Hip Ratio | 266,568 | 57 ± 17 | 103,033  (39%) | 160604  (60%) | 2931  (1%) | -0.00042 (-0.0007 to -0.0001) | 51.5 | -0.0002 (-0.0006 to 0.00005) | NS |

**Supplemental Table 6.** SGLT2 inhibitor genetic instrument characteristics

| **Chromosome** | **Position** | **Reference Allele** | **Alternative Allele** | **Beta** | **Minor Allele Frequency** |
| --- | --- | --- | --- | --- | --- |
| 16 | 31321334 | C | T | -0.00673 | 0.0436 |
| 16 | 31321569 | T | C | -0.00825 | 0.0242 |
| 16 | 31331287 | C | T | -0.00414 | 0.0632 |
| 16 | 31332332 | A | G | -0.00350 | 0.2822 |
| 16 | 31332349 | C | G | -0.0380 | 0.0146 |
| 16 | 31332634 | T | TTTTAC | -0.0024 | 0.1573 |
| 16 | 31355997 | T | C | -0.0032 | 0.0934 |
| 16 | 31357495 | C | G | -0.0024 | 0.1552 |
| 16 | 31371143 | G | C | -0.0075 | 0.0282 |
| 16 | 31373327 | T | C | -0.0102 | 0.0556 |
| 16 | 31377034 | G | A | -0.3628 | 0.0010 |
| 16 | 31381987 | A | G | -0.0029 | 0.2386 |
| 16 | 31382228 | CT | C | -0.0026 | 0.2980 |
| 16 | 31382858 | G | A | -0.0012 | 0.3083 |
| 16 | 31394352 | C | T | -0.0345 | 0.0265 |
| 16 | 31407548 | G | A | -0.0017 | 0.0430 |
| 16 | 31407654 | T | C | -0.0154 | 0.0158 |
| 16 | 31411108 | T | C | -0.0037 | 0.0629 |
| 16 | 31433872 | G | T | -0.0102 | 0.0133 |
| 16 | 31434271 | T | G | -0.0076 | 0.1952 |
| 16 | 31434472 | CT | C | -0.0011 | 0.4160 |
| 16 | 31434472 | C | CTT | -0.0044 | 0.0348 |
| 16 | 31458221 | G | A | -0.0031 | 0.0033 |
| 16 | 31459219 | T | C | -0.0026 | 0.2675 |
| 16 | 31459565 | T | A | -0.0014 | 0.0487 |
| 16 | 31464865 | C | G | -0.0016 | 0.0666 |
| 16 | 31465594 | T | C | -0.0030 | 0.0246 |
| 16 | 31477712 | C | T | -0.0111 | 0.2490 |
| 16 | 31528606 | T | A | -0.0217 | 0.0055 |
| 16 | 31528613 | T | G | -0.0019 | 0.2113 |
| 16 | 31528709 | T | C | -0.0025 | 0.0893 |

**Supplemental Table 7.** GLP-1R agonist genetic instrument characteristics.

| **Chromosome** | **Position** | **Reference Allele** | **Alternative Allele** | **Beta** | **Minor Allele Frequency** |
| --- | --- | --- | --- | --- | --- |
| 6 | 38857553 | C | T | -0.0022 | 0.4351 |
| 6 | 38907976 | A | G | -0.0069 | 0.0541 |
| 6 | 38951533 | A | G | -0.0029 | 0.1422 |
| 6 | 38984222 | C | T | -0.0045 | 0.1270 |
| 6 | 38990077 | A | G | -0.0026 | 0.4042 |
| 6 | 39048860 | C | T | -0.0145 | 0.2924 |
| 6 | 39065819 | G | A | -0.0905 | 0.0146 |
| 6 | 39072878 | A | C | -0.0157 | 0.4781 |
| 6 | 39080715 | C | A | -0.0208 | 0.4950 |
| 6 | 39086166 | G | GACAC | -0.0069 | 0.2156 |
| 6 | 39086194 | CACACACACACAT | C | -0.0447 | 0.0141 |
| 6 | 39086362 | T | G | -0.0149 | 0.0695 |
| 6 | 39086813 | C | G | -0.0003 | 0.0876 |
| 6 | 39087236 | G | T | -0.0044 | 0.0698 |
| 6 | 39087645 | A | T | -0.0082 | 0.1306 |
| 6 | 39087914 | G | A | -0.0047 | 0.4582 |
| 6 | 39104258 | G | A | -0.0052 | 0.2705 |
| 6 | 39104905 | A | G | -0.0117 | 0.3902 |
| 6 | 39190005 | A | C | -0.0070 | 0.4669 |
| 6 | 39190089 | A | G | -0.0026 | 0.0408 |
| 6 | 39190396 | A | G | -0.0003 | 0.0356 |
| 6 | 39191631 | A | G | -0.0214 | 0.0128 |
| 6 | 39229341 | G | A | -0.0052 | 0.0107 |

**Supplemental Table 8**. Multiplicative interaction analysis

| **Exposure** | **Outcome** | **OR (95% CI)** | | **P-Interaction** |
| --- | --- | --- | --- | --- |
|  |  | **SLC5A2 > Median** | **SLC5A2 < Median** |  |
| GLP1R | CAD | 0.98 (0.96-1.00) | 0.98 (0.96-1.00) | 0.92 |
| GLP1R | HF | 0.98 (0.96-1.01) | 0.97 (0.95-0.99) | 0.61 |
| GLP1R | CKD | 0.98 (0.96-1.01) | 0.95 (0.93-0.97) | 0.032 |
|  |  | **GLP1R > Median** | **GLP1R < Median** |  |
| SLC5A2 | CAD | 0.98 (0.96-0.99) | 0.99 (0.97-1.02) | 0.18 |
| SLC5A2 | HF | 0.98 (0.95-0.99) | 0.98 (0.96-1.00) | 0.79 |
| SLC5A2 | CKD | 0.99 (0.98-1.02) | 0.97 (0.96-0.99) | 0.079 |

**Supplemental Table 9.** SGLT2 inhibitor and GLP-1R agonist genetic instrument phenome-wide association study results

Caption: The P-values were adjusted according to the Bonferroni procedure.

CI = confidence interval; OR = odds ratio

| **Instrument** | **Phecode** | **Phecode description** | **OR (95% CI)** | **Adjusted P-Value** |
| --- | --- | --- | --- | --- |
| SGLT2 Inhibitor | 250.3 | Insulin pumper user | 0.96 (0.95-0.97) | <0.001 |
| SGLT2 Inhibitor | 318.0 | Tobacco use disorder | 0.97 (0.96-0.98) | 0.009 |
| GLP-1R Agonist | 250.0 | Diabetes mellitus | 0.95 (0.94-0.96) | <0.001 |
| GLP-1R Agonist | 250.2 | Type 2 diabetes | 0.95 (0.94-0.96) | <0.001 |
| GLP-1R Agonist | 250.3 | Insulin pump user | 0.94 (0.92-0.96) | <0.001 |
| GLP-1R Agonist | 276.1 | Electrolyte imbalance | 0.96 (0.95-0.98) | 0.010 |
